# A systematic review of gene editing clinical trials

**DOI:** 10.1101/2022.11.24.22282599

**Authors:** Sahar Fallah Akbarpoor Eshka, Mina Bahador, Mohammad Mahdi Gordan, Sara Karbasi, Zahra Mahmoudi Tabar, Mohsen Basiri

## Abstract

Gene editing technologies such as zinc finger nuclease (ZFN), transcription activator-like effector nuclease (TALEN), and clustered regularly interspaced short palindromic repeats (CRISPR) have revolutionized genetic engineering and now are being used in clinical gene therapy. We systematically reviewed gene editing clinical trials from ClinicalTrials.gov using a searching strategy that included all different gene editing technologies, followed by two rounds of independent assessment based on the inclusion and exclusion criteria, data extraction, and review of the background publications. 76 trials met our inclusion criteria including 30 studies on genetically engineered T-cell therapies for cancer, 19 studies on virus infections, and 26 studies on monogenic diseases. We have also analyzed the proportions to which different gene editing and gene delivery methods are used. We observed a growing trend of registered CRISPR-based trials indicating a raising interest in developing new therapeutic methods based on this technology. Overall, our study showed that there are promising phase-I and -II trials testing the safety and feasibility of gene editing in different clinical settings.

## Introduction

Gene editing allows precise and targeted changes in the genome of many living organisms by using genetic engineering tools to modify and rearrange the genome sequence, conduct biological research and industrial innovations, improve products and improve agricultural production, correct unwanted mutations, and provide treatment for genetic disorders, etc. Many genome editing tools have been introduced, and in the last decade, we have seen a significant increase in the acceptance and use of these tools. The achievements of gene editing have progressed beyond animals, and we are now experiencing the development of similar accomplishments in humans. Many researchers worldwide are working on gene editing to improve human life; luckily, some of these studies have progressed beyond animal tests and are now in various stages of clinical trials; however, there is much dispersion in these studies.

As shown in Figure 1, we first found 201 clinical trials about gene editing or genome editing tools in the clinicaltrials.gov database in this study. And after further reviewing the information on these clinical trials, we concluded that gene editing was done with the help of genetic engineering tools in only 76 studies, of which 30 studies were related to genetically engineered T-cell therapies for cancer, 19 studies related to virus infections, 26 studies are related to monogenic diseases, 21 of which are related to monogenic blood diseases.

**Figure 1:**
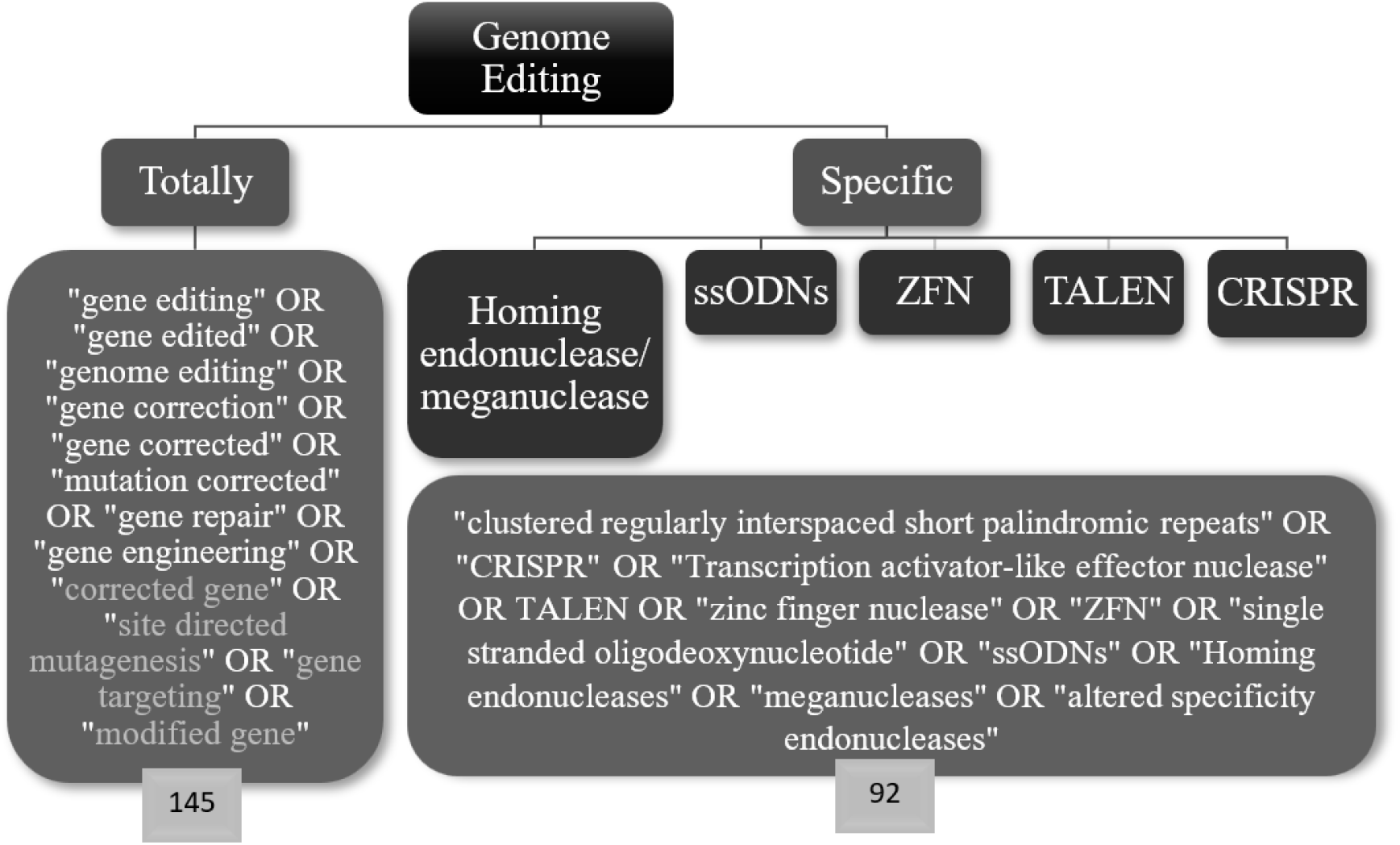
Clinicaltrials.gov database search terms. A more general keyword that we searched in the Clinicaltrials.gov database yielded 145 trials, and a more specific keyword for gene editing, which includes all genetic engineering tools, returned 92 studies. Then, all 201 studies were analyzed, and out of this number, genetic engineering tools were used for gene editing in just 76 studies.

**Figure2:**
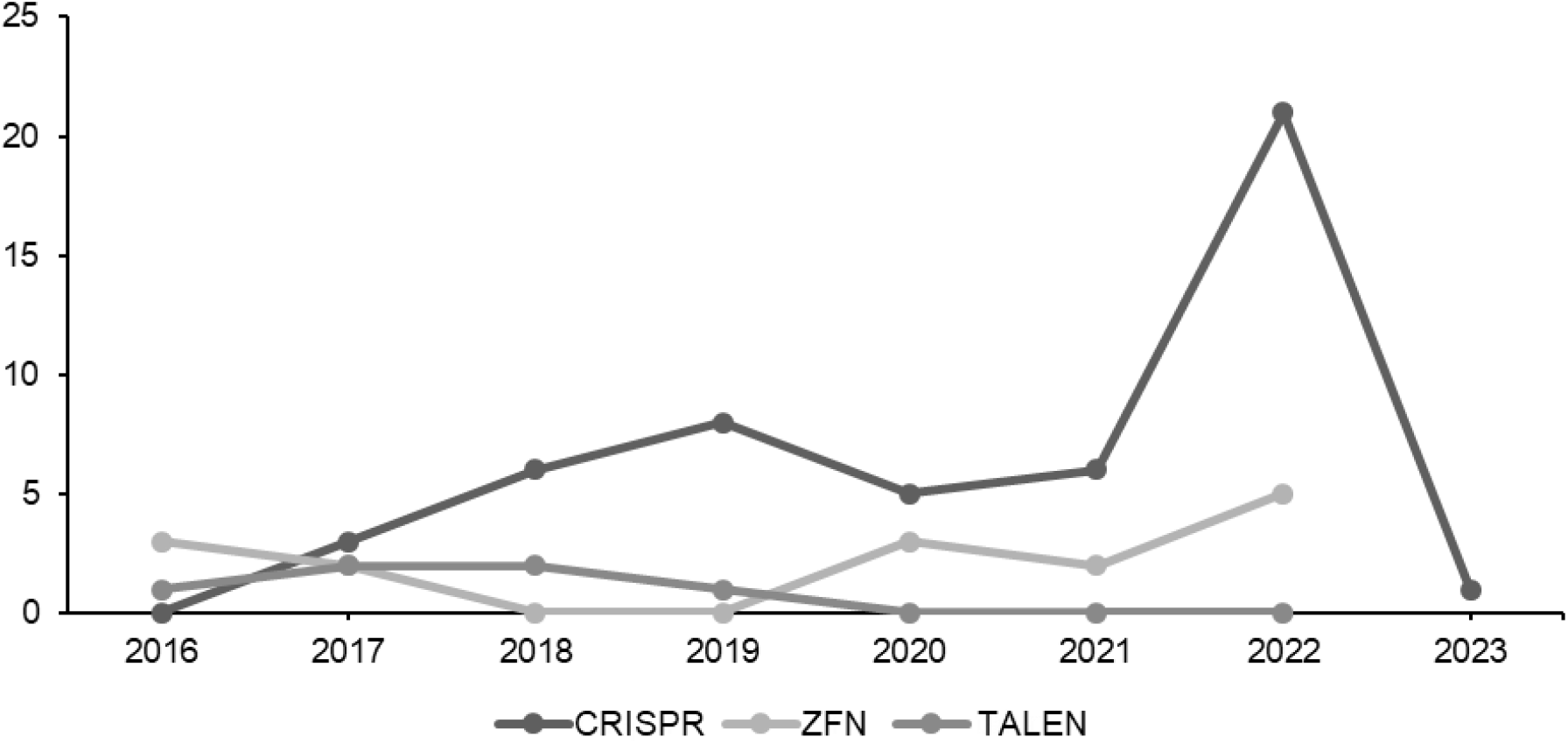
The number of clinical trials based on the year of registration and the genetic engineering tools used

**Figure 3:**
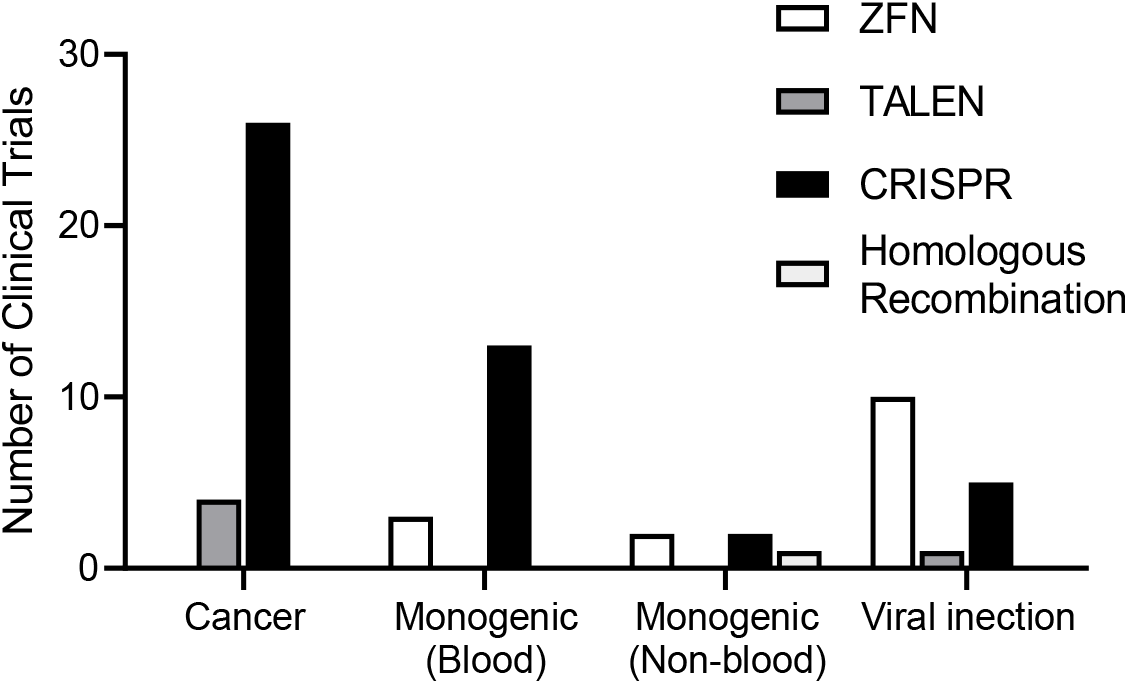
The number of clinical trials by disease and genetic engineering tools

**Figure 4:**
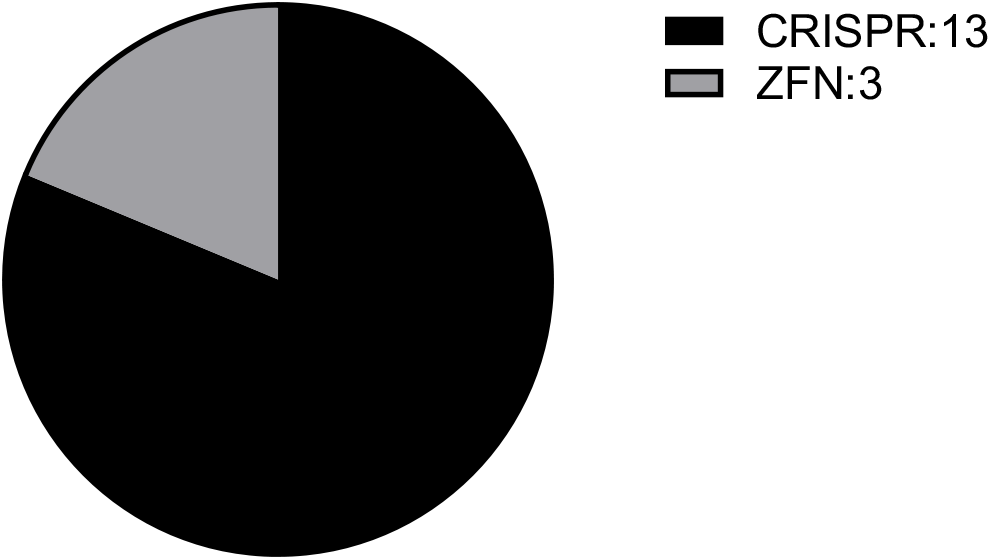
The number of Monogenic blood disorders clinical trials by genetic engineering tools.

**Figure 5:**
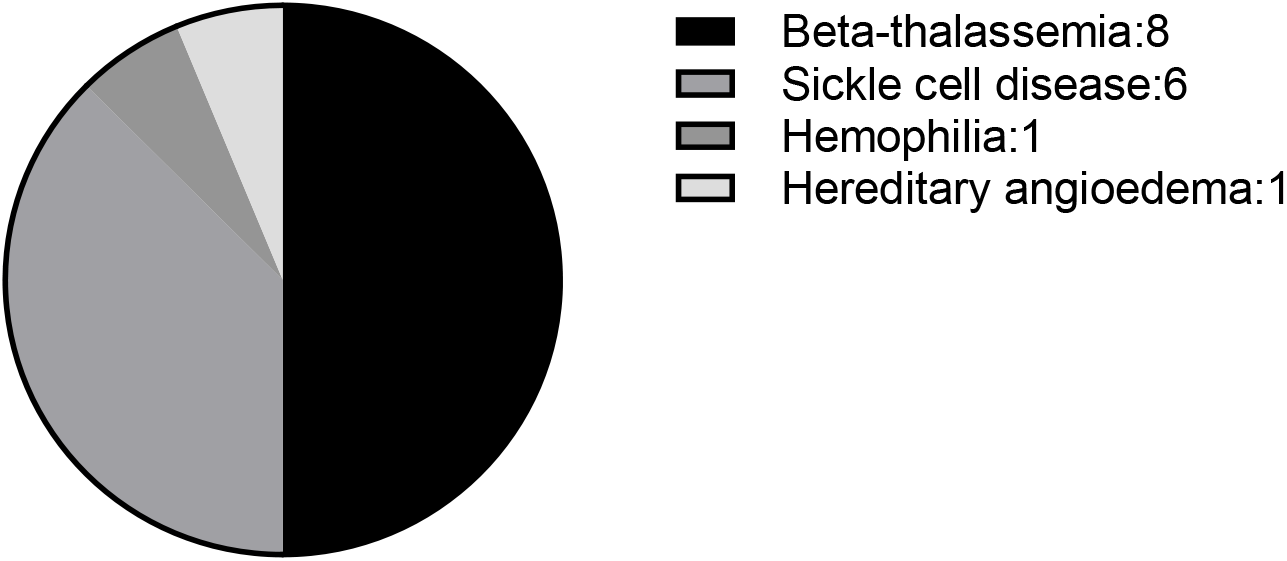
The number of Monogenic blood disorders clinical trials by the type of disease.

### Genetic engineering tools

The development of genetic engineering tools such as Clustered Regularly Interspaced Short Palindromic Repeat (CRISPR/Cas9), Transcription Activator Like Effector Nucleases (TALENs), and Zinc Finger Nucleases (ZFNs) in basic science, biotechnology, and medicine has recently increased. Four families of engineered nucleases, including engineered mega nucleases (MNs), zinc finger nucleases (ZFNs), transcription-activating nucleases (TALENs), and Cas9 protein-associated nucleases (CRISPR/CAS9), are available for gene editing (Zhang et al., 2019).

Mega nucleases, also known as homing endonucleases, identify large sequences of DNA (from 14 to 40 base pairs) and cut them (González Castro et al., 2021). Large recognition sites and low cytotoxicity compared to other methods, such as ZFN, make MNS a suitable tool for genetic engineering; Also, among the limitations of mega nucleases, we can mention the limitation in the natural occurrence of mega nucleases that cannot adequately cover the entire genome, complexity in re-engineering and low editing efficiency (González Castro et al., 2021).

The first generation of genome editing tools was Zinc Nuclease Finger (ZFN), which in this method of inducing DSB in DNA, requires a pair of ZFNs, each containing three to six Cys2 His2 zinc finger proteins, each of which recognizes a triple nucleotide code and the cleavage domain endonuclease is Foki (González Castro et al., 2021).

TALEN consists of a DNA-binding domain, TALE, and Fok1 nuclease domain; The DNA-binding domain is also composed of repeats, each of which is about 34 amino acids long, and each repetition identifies a nucleotide base in the target DNA sequence (Shimode et al., 2022), and also two amino acids determine the nucleotide specificity in positions 13 and 12 in these repetitions (Nemudryi et al., 2014).

ZFNs and TALENs create DSBs with a similar strategy of binding Fok1 endonuclease to DNA-binding proteins. In CRISPR/Cas9, the Cas9 endonuclease binds to the RNA that delivers it to the target DNA (Zheng et al., 2017).

Despite the many advances made by ZFN and TALEN techniques in genome editing, the Cas9/CRISPR system ushered in a new era of genome editing research. Three classes of the CRISPR system have been identified, of which type 2 (Class II) has been studied more than the others (Jinek et al., 2012). Based on the type 2 CRISPR system, by combining tracrRNA and crRNA, a single guide RNA (sg RNA) is created that can direct CAS9 to Protospacer Adjacent Motifs (PAMs) to make targeted genome changes; During cleavage of the target DNA, HNH and RuvC domains cause DSBs, where the HNH domain cuts the complementary strand and the RuvC domain cuts the non-complementary strand (Xiao et al., 2019).

All these elements cause DNA double-strand break (DSB); usually, non-homology end joining (NHEJ) and Homology directed repair (HDR) systems repair these breaks created in the genome. NHEJ is an efficient and error-prone repair system because it leads to frameshift mutations and gene knockout by random deletion or insertion of bases (indel) at the desired location. At the same time, HDR is a precise repair mechanism performed with an exogenously supplied donor repair template, often in the form of single-stranded oligodeoxynucleotides (ssODN) or plasmid DNA templates (Han et al., 2020).

### Delivery systems

A successful gene therapy strategy requires an efficient and safe delivery system to transfer the desired genetic material to the target cell. Gene transfer systems are divided into two general categories: viral-based and non-viral-based. Many viruses have mechanisms to deliver genetic material to the target cell. Still, only viruses that can provide high transfer efficiency and an advanced level of gene expression in the target cell will be necessary (Cevher et al., 2012).

Viral gene transfer systems are viruses engineered to have replication defects; these systems deliver genes to the target cell but cannot reproduce in the target cell (Cevher et al., 2012). One of the successful gene therapy systems available is viral vectors such as retroviruses, Adenoviruses (type 2 and 5), adeno-associated viruses, herpes viruses, pox viruses, human foamy virus-HFV, and lentiviruses. Retroviral vectors as one of the most common forms of gene transfer; Unlike adenoviral and lentiviral vectors, they can pass through the nuclear pores of mitotic cells and transfect dividing cells. The main limitations of these retroviral vectors are their low efficiency in vivo, immunogenicity problems, inability to transfer genes to non-dividing cells, and the risk of insertion, which could cause oncogene activation or tumor suppressor gene inactivation. Adenovirus types 2 and 5 can be used for gene delivery into dividing and non-dividing cells, and because of their low host specificity, they can be used for gene transfer to a wide range of tissues. Adenoviruses can deliver large DNA fragments (up to 38 kb), but unlike retroviruses, because they do not integrate into the host genome, their gene expression is very short-term. Adenoviral vectors can also cause serious side effects and even death in some patients. Although adeno-associated vectors (AAV) and adenoviral vectors are similar, AAV is more immunogenic and can have long-lasting effects on the body because it integrates into a specific location on chromosome 19. The complex vector production process and limited transgene capacity are two of the key disadvantages of these vectors (up to 4.8 kb) (Wang et al., 2019).

Helper-dependent adenovirus vectors (HdAd), the most recent generation of adenovirus vectors, have fewer limitations than previous generations, including lower immunogenicity and toxicity and a low packaging capacity (8 kb). Herpes simplex virus (HSV) combines the high transmission efficiency of a gene-deleted adenoviral vector and the long-term genome integration potential of AAVs and retroviruses, showing stable transmission and limited integration sites. One of the *Retroviridae* family is called foamy viruses (FVs). FVs are a group of non-human and non-pathogenic retroviruses that have recently been developed. The potential advantages of FV vectors include a wide range of hosts, high packaging capacity, and the ability to persist in quiescent cells. Because of these properties, FVs have a unique potential for the safe and efficient delivery of multiple genes to several different cell types (Wang et al., 2019). Herpes simplex virus (HSV) can deliver transgenic DNA up to 150 kb. Due to its neuronotropic properties, it has the most significant potential for gene transfer to the nervous system, tumors, and cancer cells. As a subset of retroviruses, lentiviruses can integrate into non-dividing cells and provide 8 kilobytes of DNA sequence. Other advantages of lentiviruses include stable and long-term expression of a transgene gene, low immunogenicity, and the ability to insert more significant genes. The insertion of transgene sequences in Poxvirus vectors is different from other vector systems. It uses homologous recombination or integration in laboratory conditions (in vitro ligation) to make recombinant vaccinia virus vectors. This virus’s highly stable insertion capacity (more than 25 KB) is one of its most important advantages, but Poxvirus’s complex structure and biology have limited its use. As a herpesvirus, the Epstein-Barr virus locates itself in the host cell’s nucleus in a latent state as an extrachromosomal circular plasmid and can be used to express large pieces of DNA in target cells (Wang et al., 2019). For successful DNA delivery to the nucleus, viruses must facilitate cell-specific binding, endocytosis, diffusion from endocytic vesicles into the cytosol, transport to the cytoplasm, transfer from one end of the nuclear envelope to the other, and finally, expression of the delivered gene. Among the advantages of these systems, we can mention therapeutic genes’ constant expression. Still, these systems have limitations, such as the use of viruses in production, immunogenicity, toxicity, and lack of optimization in large-scale production (Cevher et al., 2012).

Nonviral gene transfer systems that are introduced as an alternative to virus-based systems, based on their preparation, are classified into two physical and chemical categories; In general, physical force is used for gene delivery to increase the permeability of the cell membrane, which includes microinjection, electroporation, ultrasound, gene gun, and magnetofection And in the chemical methods that have been developed to protect genetic therapeutic materials and enable more efficient delivery, natural or artificial carriers that include polymers, liposomes, dendrimers, and cationic lipid systems are used to transfer the gene to the target cell. Compared to viral delivery systems, nonviral vectors are less toxic, less immunogenic, easier to produce, and have the potential for repeated administration. However, they are still less effective and can rarely induce transgene expression at therapeutic levels (Oliveira et al., 2017).

This study examines all the clinical trials in which gene editing has been performed; In 73% of these studies, nonviral delivery system methods include electroporation and nanoparticles. In 27% of studies, viral delivery system methods include adeno-associated viral (AAV) vector, adenoviral vector, and lentivirus vector.

### Cancer and genetically engineered T cell therapies

Individual human tumors develop due to genetic and epigenetic modifications that promote immortality while producing foreign antigens, neoantigens, that should enable neoplastic cells to be recognized by the immune system and targeted for elimination (Farkona et al., 2016). Utilizing several immunotherapy approaches such as vaccinations, oncolytic viruses, adaptive cell treatment using T cells (ACT), inhibiting active immunological suppressive pathways, or a combination of methods to strengthen and enhance the immune system to treat cancer among patients (Miliotou & Papadopoulou, 2018).

Adaptive cell therapy by T cells has been considered among the many immunotherapy techniques used to treat acute cancers (Depil et al., 2020). T lymphocytes are specially manipulated in a laboratory environment during this type of treatment and injected into the patient. Considering that the antigenic specificity of T cells is the main reason for their use in immunotherapy, T cells can be manipulated to create the ability to detect tumor antigens to remove tumor cells with the help of genetic engineering approaches to express T Cell Receptors and Chimeric Antigen Receptors (Liu et al., 2021; Miliotou & Papadopoulou, 2018). Only one injection of the anti-tumor product is required in this type of treatment. Furthermore, their long-term stability and active presence in the human body are significant. All of these factors, as well as preventing disease recurrence by identifying and destroying regrown cancer cells, can demonstrate this treatment method’s most obvious advantage and superiority over others (Galluzzi & Martin, 2017; Perales et al., 2018).

In the first trials on T lymphocytes, after deriving lymphocytes from PBMC, specific genes are knocked out to increase and improve lymphocyte activity. For example, studies using the “CRISPR/Cas9” technique on Metastatic Non-small Cell Lung Cancer (Lu et al., 2020), (NCT02793856), Advanced Hepatocellular Carcinoma (NCT04417764), Muscle-invasive Bladder Cancer (NCT02863913), Castration-Resistant Prostate Cancer (NCT02867345), Metastatic Renal Cell Carcinoma (NCT02867332), and Advanced Esophageal Cancer (NCT03081715), PDCD1 gene has been knocked out.

In the following steps, T cells infiltrating tumor tissue (TIL) against the Neo Antigens are utilized to carry out genetic manipulations. The only trial registered is for Gastro-Intestinal (GI) cancer (NCT04426669), and the CISH gene has been knocked out (Arthofer et al., 2021; Palmer et al., 2022).

With advancements in this field, techniques based on TCR and CAR were developed to produce more specific T cells. Each of these two approaches has advantages and disadvantages as compared to the other, resulting in the development of diverse applications. For more information, you may refer to the related research papers (Anikeeva et al., 2021; Chandran & Klebanoff, 2019; He et al., 2019; Morton et al., 2020; Watanabe & Nishikawa, 2021; Zhao & Cao, 2019; Q. Zhao et al., 2021). Following that, we will look at trials in this field depending on the patient’s T cells.

TRAC (TCR), TRBC (TCR), and PDCD1 (PD-1) genes were disrupted (removed, deleted) in a trial for Multiple Myeloma, Melanoma, Synovial Sarcoma Myxoid, Round Cell Liposarcoma (NCT03399448), and the NY-ESO-1 TCR gene enters T cells via lentiviruses. CRISPR/Cas9 technology was used in this research (Stadtmauer et al., 2020). During the trial related to Acute Myeloid Leukemia (NCT05066165), TRAC and TRBC genes are knocked out in T cells using CRISPR technology. It should be highlighted that TRAC is knocked out by introducing WT1-targeting TCR (a particular TCR against the antigen present on the surface of the tumor) into this gene locus (Cossette et al., 2021).

In clinical trial NCT04037566 the MAP4K1 gene was knocked out in the first research, and the Anti-CD-19 CAR gene was introduced through a lentiviral vector (Si et al., 2020). In another trial (NCT03747965) the PDCD-1 gene was knocked out, and anti-mesothelin CAR was inserted (Z. Wang et al., 2020; Wang et al., 2021). In the third study (NCT04976218), the TGFR2 gene was knocked out, and anti-EGFR was introduced using a lentiviral vector (Tang et al., 2020).

In the CRISPR/Cas9-based study for relapsed/refractory B-cell lymphoma (NCT04213469), the Anti-CD-19 gene entered the PD1 gene locus and made it inactive (Zhang et al., 2020). In the Relapsed/Refractory CD5+ Hematopoietic Malignancies study (NCT04767308) using CRISPR/Cas9 technology, the endogenous CD5 gene was knocked out, and anti-CD5 CAR was also introduced to T cells (Dai et al., 2021).

There are limitations to autologous T cell therapy, in which the patient’s lymphocytes are used to produce an anti-tumor product, including the long production process, treatment delay, risk of failure, and inability to make the maximum number of effective engineered T cells, the patient’s lymphocytes being inappropriate because of the side effects of previous treatments (chemotherapy, radiotherapy). Finally, high costs that resulted from the positive initial response and registration of numerous clinical trials efficacy measurements of the allogenic method with the approach of using lymphocytes donated from healthy people and removing the limitations can be considered by researchers as a new and effective treatment option (Benjamin et al., 2020; Depil et al., 2020). However, the allogeneic technique faces two fundamental immunological challenges: graft-versus-host disease (GVHD) and genetically modified T cells that are foreign to the host’s immune system and must be removed (allorejection). Targeting TCR constant (TRAC) or TCR constant 1 (TRBC1) and TCR constant 2 (TRBC2) genes with the aid of genome editing tools like CRISPR (Eyquem et al., 2017; Georgiadis et al., 2018)-ZFN (Provasi et al., 2012; Torikai et al., 2012)-TALEN (Berdien et al., 2014; Sommer et al., 2019) is one of the most significant and comprehensive measures to avoid GVHD. Since only one gene locus (TRAC) is responsible for the center of the constant strand α, this approach is particularly efficient in preventing the expression and positioning of TCR on the surface of T cells equipped with CAR (Osborn et al., 2016; Wiebking et al., 2021).

To avoid allorejection, beta-2-microglobulin (B2M) and class II major histocompatibility complex transactivator (CIITA) genes were utilized to target and inhibit MHC class I and II molecules, respectively (Holling et al., 2002; Krawczyk et al., 2004; Wang et al., 2015).

The trials submitted in these two areas will then be discussed.:

- This section looks into two trials using the “Talen” technique. The anti-CD123 gene was inserted into T cells in both of these researches after the TRAC gene locus was knocked out. Relapsed/Refractory Acute Myeloid Leukemia (NCT03190278) and Adverse Genetic Risk Acute Myeloid Leukemia (NCT04106076) are the two subfields in which these investigations have been done (Sugita et al., 2022).
- All trials in this section have been done with the “CRISPR/Cas9” technique, and the TRAC gene has been knocked out by inserting a gene in its gene locus. In the study related to Relapsed or Refractory T or B Cell Malignancies (NCT04502446) and Relapsed or Refractory Renal Cell Carcinoma (NCT04438083), the anti-CD70 gene (CRISPR Therapeutics, 2022a, 2022b, 2022d), in the trial associated with Relapsed or Refractory B-Cell Malignancies (NCT04035434), the anti-CD19 gene (CRISPR Therapeutics, 2022b, 2022d), in the trial of Relapsed or Refractory Multiple Myeloma (NCT04244656) the anti-TNFRSF17 (CRISPR Therapeutics, 2022d, 2022e) and finally, in the study related to Relapsed/Refractory B Cell Non-Hodgkin Lymphoma (NCT04637763), anti CD19 gene (Caribou Biosciences, 2022; Nastoupil et al., 2022), were the inserted genes in this locus. Meanwhile, in all these T cells, except for the last one [in which the PD1 gene has been knocked out], the B2M gene has been knocked out. {NCT04502446- NCT04438083- NCT04035434- NCT04244656- NCT04637763}
- The TRAC gene has been knocked out; depending on the trial; other genes have also been knocked out. The CS1 gene in a trial of Relapsed/Refractory Multiple Myeloma cancer (NCT04142619) that progressed with the “Talen” technique (Galetto et al., 2015; Galetto et al., 2016; Mathur et al., 2018); the CD52 gene in a trial of Relapsed or Refractory CD22+ B-cell Acute Lymphoblastic Leukemia (NCT04150497) that progressed with the “Talen” technique (Depil et al., 2020; Jain et al., 2020; Wells et al., 2017); and the CD52 gene in a B-cell Acute Lymphoblastic Leukemia cancer study (NCT04557436) that used CRISPR/Cas9 (Georgiadis et al., 2018; Ottaviano et al., 2021), have been knocked out. In addition, anti-CS1, anti-CD22, and anti-CD19 genes were inserted. {NCT04557436-NCT04142619- NCT04150497}
- In the two studies on relapsed or refractory CD19+ leukemia and lymphoma, both of which used the “CRISPR/Cas9” technology (NCT05037669, NCT03166878). TRAC, B2M, and CIITA genes were disrupted in the first research, and TRAC and B2M genes were disrupted in the second research (Depil et al., 2020). In both trials, the antiCD19 gene was inserted into the gene locus. {NCT05037669- NCT03166878}
- The TRAC and PD1 genes were disrupted using the “CRISPR/Cas9” technology in the Mesothelin Positive Multiple Solid Tumors study (NCT03545815), and the anti-MSLN gene was also inserted (Wang et al., 2021).
- To identify and target tumor cells missing CD19 in Relapsed or Refractory Leukemia and Lymphoma (NCT03398967), combined anti-CD19 and anti-CD20 or anti-CD19 and anti-CD22 CAR T cells were designed in this trial, and the TRAC gene was also knocked out using CRISPR/Cas9 technology (Depil et al., 2020).
- The genome of T cells has been modified using CRISPR technology in the trial for T Cell Malignancies with the reference number “NCT05397184” using BE CAR-7. Beyond what has already been mentioned, no additional information has been disclosed.

#### Modified hematopoietic stem cell (HPSC) transplantation

Patients who received the drug VOR33 in the trial coded “NCT05309733” were examined and monitored for 15 years. Blood is obtained from qualified, healthy people during treatment, CD34+ cells are isolated, and the CD33 gene is knocked out using CRISPR technology. Finally, after the cells have been modified, they are transplanted into the patient, who can then easily undergo treatments that target the CD33 marker (Hazelbaker et al., 2021; Humbert et al., 2019; Lydeard et al., 2021).

### Monogenic blood disorders

Defects in the composition or quantity of hemoglobin production result in hemoglobinopathies. Hemoglobin comprises two alpha-like chains (The gene is found on chromosome 16) and two beta-like chains (The gene is found on chromosome 11). Adult hemoglobin has two beta chains and two alpha chains, whereas fetal hemoglobin, which has a slightly different structure, has two alpha chains and two gamma chains.

#### Sickle cell disease (SCD) and thalassemia

An abnormal beta chain disorder, which can sometimes result from a mutation in the beta-globin locus, can lead to either abnormal or insufficient hemoglobin production, resulting in either thalassemia or Sickle cell disease (SCD).

#### Conventional treatments for thalassemia

Thalassemia treatment can differ depending on the severity, type, and patient. Blood transfusion is one treatment method, and people with major thalassemia, in particular, require regular blood transfusions because their bodies produce only a small amount of hemoglobin on their own. In addition, many people with beta-thalassemia receive folic acid (Vitamin B).

#### Conventional treatments for SDS

Sickle cell anemia is an autosomal recessive disease caused by a single nucleotide change in codon 6 of HBB from adenine to thymine, which results in glutamic acid instead of valine (Vakulskas et al., 2018) (Figure 6). SCD is a disease caused by a mutation in the beta-globin locus, which causes the blood cells to become stiff and sticky. People with SCD usually show symptoms from the age of 5 years. Among the complications of this disease, we can mention infection, stroke, pain episodes, and Acute Chest Syndrome (ACS). However, symptoms vary from person to person and may change over time.

**Figure 6:**
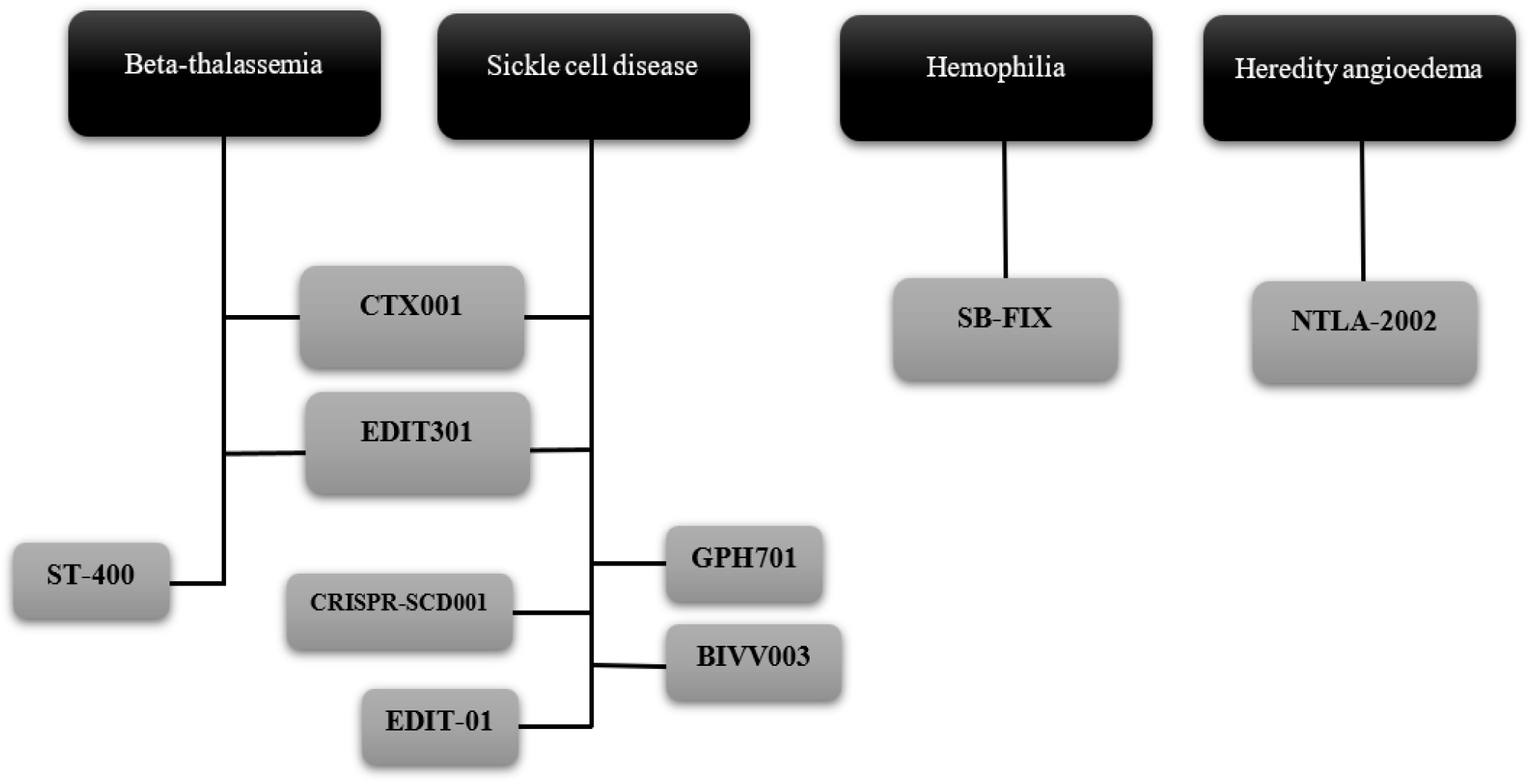
Drugs types in monogenic blood disorders

**Figure 7:**
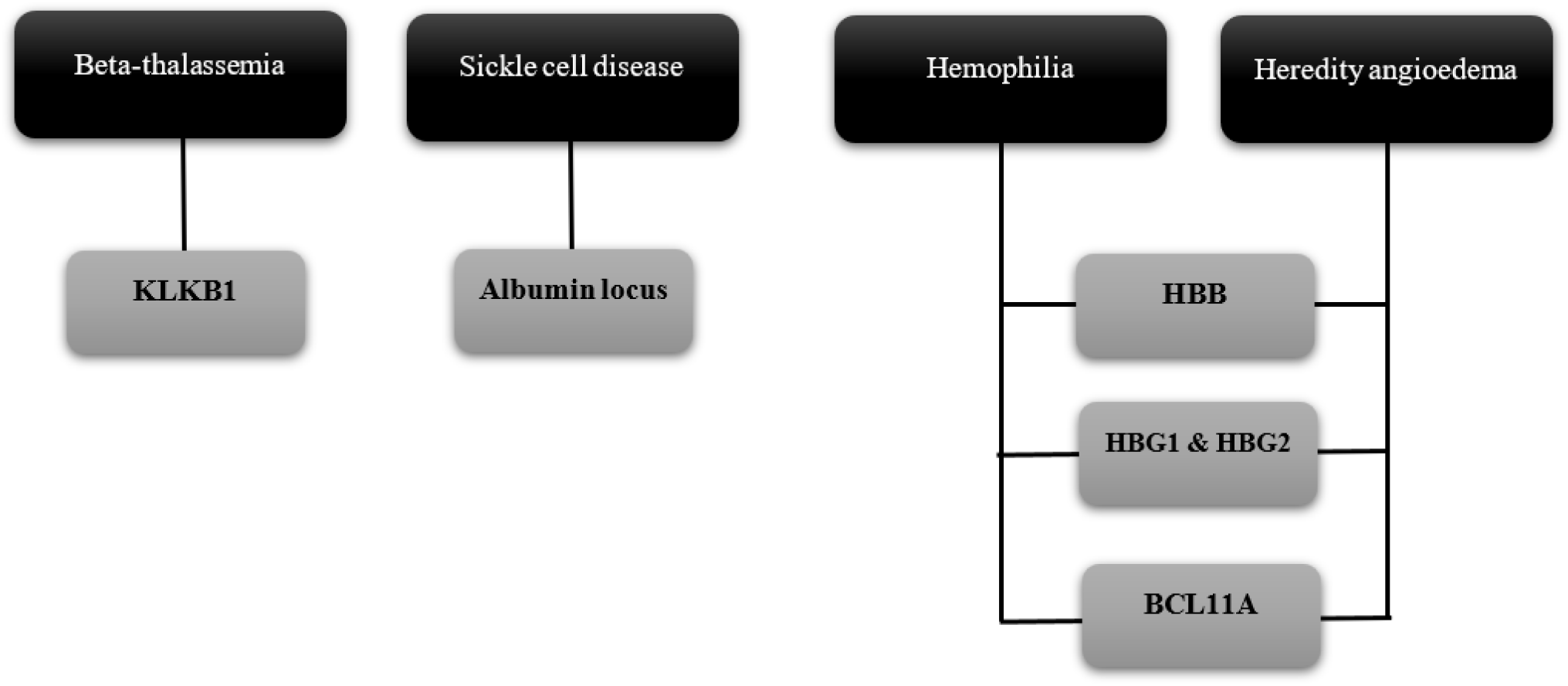
Target genes in monogenic blood disorders

The conventional cure for SCD is another person’s bone marrow or stem cell transplantation. The best bone marrow donor with stem cells is the patient’s sibling. However, this treatment is usually used for children with severe SCD because this method of treatment can be dangerous. In the following, we will discuss the series of trials registered in the treatment of hemoglobinopathy diseases (mainly thalassemia and SCD) with the help of genome editing techniques. In this article, we separate clinical trials related to thalassemia and sickle cell anemia based on the target gene:

#### HBB gene

Modifying the HBB gene changes the structure of beta chains, and when beta chains are placed next to alpha chains, hemoglobin can establish the correct form. Because mutations in the HBB gene can cause both thalassemia diseases and sickle cell disease, in all four studies NCT03728322, NCT04774536, NCT04205435, and NCT04819841, clinical trials using the CRISPR/CAS9 method focus on the HBB gene. (Studies NCT04774536 and NCT04819841 are for the treatment of SCD, while studies NCT03728322 and NCT04205435 are for the treatment of thalassemia.). The studies NCT04819841 and NCT04774536 for the treatment of sickle cell anemia use the CRISPR technique to target the HBB gene and electroporation to transfer RNP.

GPH101 is designed in such a way that a cut is created with the help of CRISPR CAS9, then a correct copy of the gene enters the target cells with the help of AAV, and the mutated part of the gene is replaced with the correct sequence by the HR mechanism. Finally, it results in the proper production of HbA (Kesavan, 2021). The participants are under the influence of Plerixafor for mobilization and apheresis before the treatment (Kanter et al., 2021).

The GPH101 ribonucleoprotein combination contains sgRNA and HiFi Cas9-R691A (high-fidelity Cas9 mutant), and HiFi Cas9 reduces OTEs (Off Target Effect). Using a correct homologous gene model allows us to make changes with greater precision, both at the nucleotide level and in the large dimensions of a gene (Vakulskas et al., 2018). The primary goal of studies NCT03728322 and NCT04205435 is to modify the HBB gene in the patient’s iHSCs, and HSCs using CRISPR/Cas and then transfer them back to the patient’s body.

#### BCL11A gene

Another way to deal with sickle cell disease is to disrupt the expression of the BCL11A transcription factor, leading to increased HbF production (Frangoul et al., 2021). Several trials (NCT03745287, NCT03653247, NCT03432364, NCT03655678, NCT04925206, and NCT04211480) are aimed at disrupting BCL11A. Studies NCT03745287 and NCT03653247 are related to sickle cell disease, and studies NCT04211480, NCT03432364, NCT03745287, and NCT04925206 are related to thalassemia patients. All of these studies aim to increase fetal hemoglobin production by reactivating gamma-globin.

Producing more fetal-type hemoglobin (HbF; α2γ2) is one of the therapeutic solutions for thalassemia and, of course, SCD. This therapeutic solution helps produce fetal hemoglobin instead of adult hemoglobin (HbA; α2β2). And, because beta globulin mutations cause both thalassemia and SCD, not relying on the body to produce beta globulin simultaneously can be a treatment for both thalassemia and SCD patients. (Reduces HbS polymerization). Reactivating the HBG gene can be accomplished in two ways: either by downregulating the expression of the HbF production inhibitory gene, such as BCL11a, or removing the cis-regulatory elements of the HBG1/2 promoter region (L. Wang et al., 2020). By targeting the BCL11a gene and removing its erythroid enhancer, the inhibition of this gene from producing the gamma gene was reduced, and HbG expression increased as a result.

ST-400 is the individual’s autologous CD34+ hematopoietic stem/progenitor cells, into which the mRNA encoding ZFN was electroporated under ex vivo conditions. ZFN is produced, and the GATA motif in the BCL11a gene’s ESE is influenced by INDEL (Walters et al., 2021). This research group previously reported that plerixafor and a combination of granulocyte colony-stimulating factor (G-CSF) were used to dual mobilize HSPCs. BIVV003, an ST-400 analog drug, is used for SCD patients in the NCT03653247 study. However, the use of G-CSF in SCD patients is not recommended due to the risk of clinical complications. Only plerixafor is thus used to mobilize HSPCs.Before this clinical procedure, BIVV003 was injected into immune-deficient NBSGW mice, which causes strong and long-lasting engraftment but does not change the number of HSPC or their progeny differentiation (Moran et al., 2018). Both the NCT04925206 and NCT04211480 studies use CRISPR to target the BCL11A gene INDEL, and they do by electroporation and inserting RNP into hHSPC cells. CTX001 is one of the most extensively studied drugs. CTX001 are CD34+ human autologous hematopoietic Stem and Progenitor Cells that have been treated with CRISPR CAS9 in their BCL11A gene enhancer INDEL. Finally, CTX001 reactivates the production of fetal hemoglobin.

NCT03655678, NCT05356195, and NCT05477563 are studies that investigated the efficacy and safety of this drug in patients with blood transfusion-dependent thalassemia. NCT03745287 and NCT05329649 studies looked into the safety and efficacy of the same drug on sickle cell anemia patients, respectively. CTX001 is being studied for both thalassemia and sickle cell anemia.

Monitoring all 114 patients who received CTX001 is the focus of the NCT04208529 study. According to preliminary results published on November 5, 2020, all seven participating patients (5 with TDT and 2 with sickle cell anemia) had an overall increase in Hb and HbF levels. Blood transfusions were stopped in TDT patients quickly after receiving CTX001, and the first patient with TDT who received the medicine was 15 months without the need for a blood transfusion. After receiving CTX001, patients with SCD did not experience any VOC (Vaso-occlusive crisis). The first SCD patient received the medication without a VOC for an entire year. Only one of the TDT patients had four inexplicable symptoms that CTX001 could have caused: headache, hemophagocytic lymphohistiocytosis (HLH), acute respiratory distress syndrome, and idiopathic pneumonia syndrome. All of these symptoms occurred as a result of HLH. There have been no other SAEs (Serious Adverse Events) associated with CTX001 reported in other patients with TDT or SCD (Frangoul et al., 2020).

#### HBG1 and HBG2 promoter

EDIT-301 is a cell therapy treatment that uses autologous CD34+ cells. Cas9a RNP edits CD34+ cells to increase the expression of gamma-globin, and instead of generating an incorrect version of ADULT hemoglobin, the amount of HbF in the blood increases, resulting in SCD treatment (NCT04853576) (De Dreuzy et al., 2020). EDIT-301, like CTX001, has been studied for the treatment of SCD as well as blood transfusion-related beta-thalassemia. NCT05444894 investigates the efficacy of EDIT-301 in patients with transfusion-related beta-thalassemia, while NCT04853576 investigates the same drug’s effect on SCD patients. Previous trials, which formed the basis for the NCT04853576 trial, aimed to determine whether ingesting SpCas9-based RNP or Sp12a-based RNP (or Cpf1) would be more effective. The results of their study on NBSGW mice showed that Cas12a could create larger deletions and higher frequencies of productive indels than SpCas9 RNP. As a result, this study identified a particular Cas12a RNP that efficiently edited the distal region of CCAAT in the HBG1/2 promoter in CD34+ cells in NBSGW mice, leading to the stability of productive indels and elevated HbF expression (De Dreuzy et al., 2019).

The effectiveness of Cis-regulatory elements in the beta-globin locus, primarily focused on gamma-globin promoters or reducing BCL11a expression in erythroid cells, is the subject of another study. The results showed that the HBG1/2 promoter edit was highly specific and did not cause off-target electroporation, resulting in more stable HbF expression in the erythroid lineage. Cells having the BCL11a gene altered, on the other hand, did not differentiate or survive (Chang et al., 2018). BEAM 101 is the first candidate for patient-specific autologous cell therapy with a basic editing approach for hemoglobinopathy, which has been used to compensate for hemoglobin deficiency and potentially reduce sickle cell disease symptoms (Karen O’Hanlon Cohrt, 2021).

Base editors are derived from CRISPR Cas technology as engineered fusion enzymes that can modify a cytidine base (cytosine base editor, CBE) or adenosine (adenine base editor, ABE) in genomic DNA without causing a double-stranded breakdown. Ex vivo electroporation delivers BEAM 101 baseline editing reagents to patient-derived hematopoietic stem cells. BEAM 101 is engineered with an ABE that combines edits of A → G in the promoters of the HBG1 and HBG2 genes, which regulate HbF expression, disrupting repressor connectivity and increasing gamma-globin expression (CRISPR MEDICINE News, 2022; Karen O’Hanlon Cohrt, 2021).

In the NCT05442346 research, gamma globin is reactivated in autologous hematopoietic stem cells under ex vivo conditions employing Glycosylase Base Editors. These glycosylase base editors (GBEs) are composed of a cytidine deaminase, an uracil DNA glycosylase, and a Cas9 nickase (Ung). The DNA repair process is started when Ung cleaves the deaminase-generated U base, creating an apurinic/apyrimidinic (AP) site (D. Zhao et al., 2021).

#### Hemophilia

Hemophilia is a monogenic and recessive x-linked hereditary disorder. This condition is caused by a deficiency of factor 9, which causes slow blood coagulation. The only clinical gene editing trial for this condition used SB-FIX (study NCT02695160). SB-FIX used the ZFN method, in which the gene encoding factor9 was delivered to the patient’s own hepatocytes’ Locus albumin using AAV2/6.

#### Hereditary Angioedema (HAE)-3

An uncommon genetic illness known as hereditary angioedema (HAE) is responsible for severe, sporadic, and unexpected inflammatory attacks that affect different body organs and tissues. These attacks may be painful, damaging, or even deadly. One in 50,000 persons is affected with HAE, and the current therapies typically include lifetime treatments that may require chronic intravenous (IV) or subcutaneous (SC) injections; usually, twice a week or daily oral administration may be required. However, there is still the possibility of unexpected episodes (Intellia Therapeutics, 2021).

The NCT05120830 study looks at NTLA 2002’s safety, tolerability, activity, pharmacokinetics, and pharmacodynamics to permanently reduce plasma kallikrein activity and, thus, the severity of HAE attacks. NTLA 2002 employs the CRISPR/Cas 9 gene editing tool, packaged in lipid nanoparticles and absorbed by liver cells after an intravenous dose, disrupting the activity of a gene called KLKB1. This gene encodes instructions for the synthesis of prekallikrein, a kallikrein precursor protein. In people with HAE, kallikrein levels rise, resulting in excess bradykinin, an inflammatory molecule that causes tissue swelling (Intellia Therapeutics, 2021).

### Viral infections

In the viral infections section, 19 clinical trials were examined. 13 studies are related to HIV disease (two of which were follow-up), three studies are related to HPV disease, 1 study is related to HSV disease, 1 study is related to EBV disease, and 1 study is associated with COVID disease.

#### Human Immunodeficiency Virus (HIV)

Acquired immune deficiency syndrome (AIDS) is caused by the human immunodeficiency virus (HIV), which infects immune cells (helper T cells, monocytes, and macrophages) leading to immunodeficiency and clinical symptoms in the patient. According to the statistics published by the World Health Organization (WHO) in 2021, the number of HIV-infected people worldwide was 38.4 million, of which 650,000 died (World Health Organization, 2022c). Human immunodeficiency virus type 1 (HIV-1) infection is a global health problem, and successful treatment options are still under investigation (Mohamed et al., 2021). Different drugs available in treating patients with HIV can cause various side effects. In addition, people who use the same HIV drugs may have different side effects (HIVinfo.nih.gov, 2021). Challenges facing current medications that are part of lifelong antiretroviral therapy (ART) include toxicity, the development of drug-resistant HIV-1 strains, treatment cost, and the inability to eradicate the provirus from infected cells (Mohamed et al., 2021).

According to the above-mentioned challenges, new anti-HIV-1 treatments that can prevent the progression of the disease, especially the onset of the AIDS, or eliminate it, are needed. While the development of an HIV-1 vaccine has also been challenging, recent advances suggest that it is possible to prevent infection of HIV-1-susceptible cells in HIV-1-infected individuals by targeting the chemokine receptor type 5 (CCR5) (Mohamed et al., 2021). CCR5 is the major receptor for the human immunodeficiency virus (HIV) (Tebas et al., 2014). One of the strategies that have been recently proposed for the possible treatment of HIV patients is the removal of CCR5 proteins from T cells. CD4+ T cells are one of the essential types of T cells (NCT00842634). Treatment based on targeting CCR5 generally involves gene editing techniques, including ZFN, CRISPR, blocking CCR5 using antibodies or antagonists, or a combination of both (Mohamed et al., 2021). The presence of CCR5 proteins is necessary to enter a special type of HIV called tropical CCR5 (CCR5 tropic) into immune cells, and in their absence, T cells will be immune from being infected by this type of HIV (NCT00842634).

Among the 13 existing clinical trials conducted on HIV, three studies use the CRISPR/CAS9 technology, one of which was a follow-up study in which no treatment or intervention was performed. ZFN technique was used in 10 other trials through different approaches. In 5 studies, genetic manipulation has been performed using the ZFN SB-728-T technique. In 4 studies, the ZFN SB-728mR was used, and one follow-up study reviewed the previous two techniques, which means no treatment or gene editing has occurred in it.

Except for the study NCT05144386 and its follow-up, all other 11 studies target CCR5 gene in ex vivo settings. Moreover, there is no gender restriction in these studies. On the other hand, study number NCT05144386 has limited its investigations only to the group of men and is in vivo with no information available on the targeted gene.

##### Genetic manipulation based on ZFN SB-728-T technique

The common points among these five trials are that all these studies were done in the ex vivo environment, and CD4+ T cells are targeted for genetic manipulation. The following commonality between these five studies is the genetic material used, which in all of them was the viral genome (adenovirus). Therefore, the transmission system is also common in all of them, which is an adenovirus. Meanwhile, four studies have been completed, but only one has a reported result, and its identification number is NCT01543152. Study No. NCT03666871 is the only study in this group that has not been completed yet. Although the age limit between 18 and 70 years has been applied in this study, other studies have investigated people 18 years and older. NCT00842634 and NCT01044654 have only been investigated in phase one, but the other three have advanced to phase two. The final important and noteworthy point is the gene editing done in these studies.

NCT00842634: Knock out CCR5 Protein

NCT01044654: Removing the CCR5 gene

NCT01252641: Removing the CCR5 gene

NCT01543152: Knock out CCR5 Protein

NCT03666871: Elimination of CCR5 Receptors

##### Genetic manipulation based on ZFN SB-728mR technique

The commonality of these four studies is their genetic material, which is based on mRNA, and the transfer method, which is electroporation. In addition, the CCR5 gene has been disrupted in these four studies. The target cell line in the NCT02500849 study differs from the others and is CD34+ Hematopoietic Stem/Progenitor Cells, but in the other studies, CD4+ T-cells have been treated as the target cells. NCT0250084, and NCT03617198, have not been completed yet. NCT02225665 is the only study that has been completed, and its results have been reported. Another noteworthy point about this study is that it is the only trial that has progressed to phase two, while the others have only progressed to phase one. NCT02388594 has also been completed among these four studies, but no results have been reported.

One of the studies with the identification number NCT04201782 is a follow-up study and has investigated the previous two techniques, namely Zinc Finger Nucleases SB-728-T and SB-728mR-T. No treatment was planned in this follow-up study.

##### Genetic manipulation based on CRISPR/CAS9 technique

These three studies have fundamental differences from the other ten studies. In the first trial with the identification number NCT03164135, the target cell line for manipulation was CD34+ Hematopoietic Stem/Progenitor Cells (HSPCs) genetic material used was RNPs. The transmission system is nucleofection, and the CCR5 gene is disrupted (Indel). The study subjects’ age limit was 18 to 60 years, but no gender limit was applied. No results have been published from this trial yet, and studies are still ongoing (Xu et al., 2019).

The second study, with identification number NCT05144386, is an entirely different study from all previous studies and has only two aspects in common with the others. First, as in the previous investigation, the CRISPR/CAS9 technique was used for gene editing, and second, in both, RNPs were used as genetic material, although the details of the RNPs used were also different.

EBT-101 is a unique in vivo CRISPR-based therapy designed to remove large portions of HIV proviral DNA. The experimental program uses CRISPR-Cas9 and dual-guide RNAs to target three sites in the HIV genome, thereby eliminating large sections of the HIV genome and minimizing the infectivity of the virus. EBT-101 uses adeno-associated virus (AAV) for a single intravenous infusion to functionally cure HIV infection (GlobeNewswire, 2022). For more details, you can visit the Excision BioTherapeutics website.

#### Human Papillomavirus (HPV)

Cervical cancer is the fourth most common cancer among women worldwide, with an estimated 604,000 new cases and 342,000 deaths in 2020. About 90% of new cases and deaths worldwide in 2020 occurred in low- and middle-income countries. Two types of human papillomavirus (HPV) (16 and 18) are responsible for approximately 50% of high-grade cervical precancers. HPV is mainly transmitted through sexual contact, and most people become infected with HPV shortly after starting sexual activity. More than 90% of them eventually clear the infection. Vaccination against HPV and screening and treatment of precancerous lesions is a cost-effective way to prevent cervical cancer (World Health Organization, 2022a). According to reviews conducted on the Clinicaltrials.gov website, by September 14, 2022, three studies had been found that sought to find a solution to the HPV virus using gene editing. The identification numbers of these three studies are NCT03057912, NCT02800369, and NCT03226470. All of them were conducted only on women who were in the age range of 18 to 50 years, and no case of men participated in this study. Another common point between these three studies is that the delivery system in all three studies was nanoparticles, and all of them were done in vivo. No results have been published yet, and all three studies are still ongoing.

The first study (NCT03057912) aims to investigate the safety of TALEN and CRISPR/Cas9 in the treatment of patients with malignant neoplasms associated with human papillomavirus without using any invasive method. In this trial, the HPV16/18 positive cervical cancer cell line was subjected to genetic manipulation, and E6 and E7 genes were knocked out or disrupted.

In the second study (NCT02800369), unlike the previous study, the ZFN method was used for genetic manipulation and only targeted the E7 gene. The genetic material transferred in this study is a plasmid named “pcDNA3.1 plasmid”. In this method, like the previous one, no invasive methods are used for treatment. The target cell line in this study is the same as the previous one, HPV-16/18 positive cervical cancer cells, in which only the E7 gene is disrupted.

The third study (NCT03226470) used only the TALEN method to treat these patients. It should also be noted that in this study, like the first study, E6 and E7 genes were disrupted, but the difference is that only the HPV-16 was targeted here.

#### Epstein-Barr Virus (EBV)

B-cell lymphoma disseminated by Epstein-Barr virus (EBV+DLBCL) is an aggressive malignancy that is mainly resistant to existing treatment regimens and, therefore, could be attractive for immunotherapy-based therapies (Quan et al., 2015). Only one clinical trial in the field of gene therapy for viral infections associated with the EBV virus had been registered on Clinicaltrials.gov by September 14, 2022, which is “NCT03044743”. During this trial, the PD-1 gene belonging to EBV-CTL cells was knocked out with the help of the CRISPR/Cas9 technique, and three different drugs were used to improve the conditions of this gene therapy: Fludarabine: to improve the safety of the environment; Cyclophosphamide: to improve the safety of the environment; Interleukin-2: to maintain the survival of injected T cells

In this study, using the electroporation technique, plasmids containing sgRNA and Cas9 were introduced against PD-1 into CTL cells (Su et al., 2016). There was no gender restriction in the sampling of people, but the age limit applied was from 18 to 75 years. The result of the EBV test of all the participants in this study was positive, and the experiment was not performed on healthy people.

#### Coronavirus disease (COVID-19)

Acute respiratory syndrome virus (SARS CoV-2), the causative agent of COVID-19, is an enveloped virus whose genetic material is a single-stranded RNA with positive polarity (InvivoGen, n.d.). According to the statistics published by the World Health Organization, as of June 14, 2022, 533,816,957 people were infected with this virus, and unfortunately, 6,309,633 of them died (World Health Organization, 2022d). For this reason, many efforts have been made to control this dangerous virus’s spread.

##### Previous treatment

a. Antiviral agents: There is no available evidence from randomized controlled trials (RCTs) to recommend specific anti-SARS-CoV-2 treatment for patients with suspected or confirmed infection with COVID-19. Lopinavir (LPV) inhibits the protease activity of coronavirus in vitro and animal studies. But still, these drugs cannot be mentioned as a treatment solution (Zhai et al., 2020).
b. Chloroquine and hydroxychloroquine: Chloroquine is a widely used antimalarial and autoimmune disease drug reported as a potential broad-spectrum antiviral drug (Rolain et al., 2007; Savarino et al., 2006; Yan et al., 2013; Zhai et al., 2020). Chloroquine prevents virus transmission by increasing the endosomal pH required for virus/cell fusion and interfering with the glycosylation of SARS-CoV cellular receptors. However, the point is that the optimal dose of chloroquine for SARS-CoV-2 should be evaluated in future trials because the optimal dose of this treatment has not been determined yet (Zhai et al., 2020).
c. Corticosteroids, antibodies, and vaccines can also be mentioned among other treatments, but if we can use genome editing techniques with fewer side effects, we can have a more effective treatment.

The current trial with the identification number NCT04990557 is the only study on the Clinicaltrial.gov site conducted on the treatment of the COVID-19 disease that gene editing has been performed. In this study, PD-1 and ACE2 genes were knocked out using the CRISPR/Cas9 technique, and this manipulation took place in a laboratory environment (ex vivo). The target cells in this genetic manipulation were human CD8+ T cells. The people under study in this trial were in the age range of 18 to 70 years, and gender restrictions were not applied. The purpose of this study is to check the safety of this genetic manipulation (knockout of these two genes) and to check whether this genetic change leads to longer-term immunity and, thus, in the next encounter with this virus, will the body have a proper response to re-infection and eliminate the virus or not? In this trial, people will have two treatment cycles, and in each cycle, they will be injected with three different doses of manipulated cells. Initial dose 20%, second dose 30% and third dose 50% (NCT04990557).

#### Herpes Simplex Virus (HSV)

Herpes simplex virus type 1 (HSV-1) is an enveloped virus whose genetic material is double-stranded DNA (Birkmann & Zimmermann, 2016). The primary route of HSV-1 transmission is through oral-to-oral contact causing oral and also genital herpes. The latest available estimated prevalence of HSV-1 infection in 2016 is 67% of the population with most cases HSV-1 infections are acquired during childhood (World Health Organization, 2022b).

##### Current HSV medications

Acyclovir, a guanosine analog, was one of the first antivirals to confirm excessive selectivity and low toxicity. Acyclovir is as lively in opposition to HSV-1 and HSV-2 as it is far in opposition to VZV. Acyclovir is used for the systemic remedy of HSV infections, along with genital and labial herpes, in addition to HSE, and for topical remedy of labial herpes (bloodless sores). In order to increase the oral bioavailability of acyclovir, the valine ester, valacyclovir, changed developed. Valacyclovir is indicated as a short-time period remedy for labial herpes in addition to episodic and suppressive remedies for genital herpes. Besides acyclovir and its prodrug valacyclovir, in addition, nucleoside analogs are authorized for HSV remedy: penciclovir and trifluridine. Different capsules are prescribed to deal with HSV (Birkmann & Zimmermann, 2016).

##### Limitations of current HSV drugs

Ideally, treatment should begin in the prodromal stage. This is because there is only a short window to control the virus during replication. Furthermore, nucleoside analogs (as well as other treatments such as docosanol) generally have limited efficacy and provide only modest improvements in wound healing time or episode duration. This is also true for suppressive therapy, as virus shedding and outbreaks occur during therapy, resulting in a 50% reduction in infection as well. With the above points in mind, there is room for better and more effective treatments (Birkmann & Zimmermann, 2016).

##### Vaccines

Currently, there is no commercially available vaccine against HSV infection. Prophylactic vaccination approaches have failed so far, including a phase 3 trial of the HSV-2 glycoprotein D (gD2) containing vaccine ‘Herpevac’ (GSK) involving more than 8,000 HSV-1/HSV-2 seronegative women. The most potent therapeutic vaccine candidate reported is Genosia Biosciences’ her GEN-003, a T cell-targeted immunotherapy containing the viral antigens ICP4 and gD2 with an in-licensed adjuvant (Birkmann & Zimmermann, 2016).

Trial No. NCT04560790 is the only study on the Clinicaltrials.gov website that has evaluated gene editing to combat herpes simplex virus 1 (HSV1) and investigated its safety in humans. The target tissue for this treatment was the cornea of the human eye. BD111 is a new product being investigated for gene editing in this trial. A similar treatment method has never been performed in this field. Six people from the target population of 18 to 70 years have been considered for this research, and there is no gender restriction. In this study, by means of BDmRN delivery technology, CRISPR/Cas9 mRNA was used to manipulate corneal tissue cells, and two genes, UL8 and UL29, were destroyed (NCT04560790).

### None-blood monogenic disorders

#### II (MPS I & II) & Mucopolysaccharidosis type I

Both of these conditions are lysosomal illnesses associated with the X chromosome. Mucopolysaccharidosis type I is caused by genetic changes in the IDUA gene, which results in a decrease or total loss of the -l-iduronidase (IDUA) enzyme. Heparan and dermatan sulfate molecules accumulate in affected people’s tissues when this enzyme is deficient (Clarke, 2021).

Mucopolysaccharidosis II disease is caused by the lack of lysosomal enzyme iduronate 2-sulfatase (iduronate 2-sulfatase). This causes an accumulation of glycosaminoglycans (GAGs) in the tissue of affected individuals, ultimately shortening their lives (Laoharawee et al., 2018).

The trial for MPS I disease was registered with the number “NCT02702115”, whereas the trial for MPS II disease was run under the number “NCT03041324”. The biological drug SB-318 is used in the first experiment, while the biological drug SB-913 is used in the second trial; both use the same technology, but they target different genes depending on the type of disease. ZFN technology is used throughout the treatment process with these two medications to insert the healthy gene version encoding the IDUA and IDS enzymes, respectively, and this package is inserted into the AAV-based transmission system as a viral genome. The albumin gene locus is targeted in hepatocytes, and after making a cut in this area, ZFN inserts a healthy gene locus in this spot, so the gene related to IDUA and IDS enzymes is more expressed as a result of the high expression of the albumin gene (Laoharawee et al., 2018; Ou et al., 2019; Pagant et al., 2021).

Following the first two trials, a third trial with the identifier “NCT04628871” has also been carried out; this one is a ten-year follow-up study for the biological drugs SB-318, SB-913, and SB-FIX. It looks at individuals who have received this type of treatment.

#### Heterozygous familial hypercholesterolemia (HeFH)

This disease is a single-gene disorder caused primarily by heterozygous mutations in the LDLR gene, which codes for the LDL receptor. This causes an increase in the level of LDL in the blood, which can eventually lead to cardiovascular disease (Yuan et al., 2006).

The drug VERVE-101 was used in the trial with the number “NCT05398029,” and CRISPR technology is used in this treatment method, in which CRISPR is placed in the form of RNA in a lipid nanoparticle and enters the body under in vivo conditions, targeting and knocking out the PCSK9 gene in liver cells. This function increases LDLR gene expression while decreasing blood LDL levels (Musunuru et al., 2021).

#### Leber congenital amaurosis (LCA10)

This condition is a severe form of retinal dystrophy brought on by a CEP290 gene mutation. Congenital blindness or poor vision results from this point mutation (Maeder et al., 2019).

The drug EDIT-101 was used in the disease-related trial with the identifier “NCT03872479.” This medication targets photoreceptor cells using CRISPR technology placed in an AAV5 viral vector. The technique allows the mutation to be inverted or deleted, restoring CEP290’s functional expression (Maeder et al., 2019).

##### Phenylketonuria (PKU)

Phenylketonuria is an autosomal recessive disorder of phenylalanine metabolism caused by a defect in the enzyme phenylalanine hydroxylase (PAH). In this disease, the high phenylalanine concentration causes brain dysfunction (van Spronsen et al., 2021).

HMI-103 drug has been used in the trial related to this disease with the number “NCT05222178”. The mechanism of action of the HMI-103 drug is that first, the healthy version of the gene encoding the enzyme phenylalanine hydroxylase (PAH) is inserted into the AVVHSC15 viral vector, and the liver cells are targeted. After these viruses enter the liver cells due to the homologous recombination mechanism, the healthy gene fragment is placed in the defective gene location, thus leading to the recovery of PAH enzyme function [0].

#### Metabolic disorders

The results of clinical data about islet transplantation demonstrate that beta cell replacement approaches in patients with diabetes who require insulin may have therapeutic benefits. PEC-Direct, a candidate stem cell-derived insulin-producing cell product currently being evaluated in the clinic, uses a non-immunoprotective delivery device that allows direct vascularization of the cell therapy. Although this approach has valuable potential, PEC-Direct requires long-term immunosuppression to prevent rejection since the patient’s immune system recognizes these cells as foreign. As a result, PEC-Direct is being developed as a treatment for patients with type 1 diabetes who are at high risk of complications (CRISPR Therapeutics, 2022c). CRISPR Therapeutics AG, the sponsor of this technology, claims that their gene editing technology has the potential to protect the transplanted cells from the patient’s immune system. This technology is provided by ex vivo editing of immunomodulatory genes in the stem cell line used to produce pancreatic-lineage cells. The speed, specificity, and multiplexing efficiency of CRISPR/Cas9 make this technology ideally suited to this task. Through its allogeneic CAR-T programs, the company has developed significant expertise in editing immune-evasive genes. The combination of ViaCyte’s stem cell capabilities and CRISPR Therapeutics AG’s gene-editing capabilities has the potential to enable a beta-cell replacement product that may provide lasting benefits to patients without triggering an immune response (CRISPR Therapeutics, 2022c). The result of the collaboration work of these two companies is a product called VCTX210A (NCT05210530). The combined product VCTX210A (Unit) includes two components: first, Allogeneic pancreatic endoderm cells (PEC210A) genetically modified using short palindromic repeats of cluster regular/CRISPR-associated protein 9 (CRISPR/Cas9) to enhance immune evasion and survival; second, a durable, removable, perforated device designed to deliver and maintain PEC210A cells.

## Conclusion

Gene editing technologies are being investigated in several clinical trials, paving the way for new therapies to fight cancer, infectious disease, and genetic disorders. Our systematic review of registered clinical trials showed growing interest in CRISPR technology in recent years. Not surprisingly, genetically modified T cells as a very successful cell-based gene therapy, has been a very attractive platform to exploit gene editing technologies. These efforts can potentially lead to off-the-shelf and more efficient T cell therapies for cancer and hard to treat infectious diseases. Several monogenic disorders have also been targeted using gene editing-based strategies. Although we should wait to see the effectiveness of these approaches in comparison to the conventional viral-based strategies, increasing number of gene-editing clinical trials shows the growing interest of the researchers to develop novel gene therapies based on these technologies.

## Data Availability

All data produced in the present work are contained in the manuscript.

